# Association between blood urea nitrogen-to-albumin ratio and kidney stones: A cross-sectional study based on NHANES

**DOI:** 10.64898/2026.09.16.26363211

**Authors:** Fu Zhu, Jianqiang Bian, Dongpeng Zhang, Chao Wang, Yujie Wang, Guangbin Zhu

## Abstract

**Background:** Kidney stones are a disorder in which urinary solutes precipitate to form crystalline aggregates in the urinary space. The blood urea nitrogen-to-albumin ratio (BAR), a nutrition and inflammatory indicator, has an unclear association with kidney stones.

**Methods:** Using NHANES 2007–2020 data, we compared baseline characteristics between kidney stone formers and controls and evaluated the BAR–stone relationship via weighted multivariate logistic regression (WMLR), restricted cubic splines (RCS), stratification analyses, and XGBoost, and performed an exploratory analysis in an ICU cohort, the Medical Information Mart for Intensive Care IV (MIMIC-IV).

**Results:** A total of 23,652 participants (2,318 with kidney stones) were included, and 21 baseline features differed significantly between groups (P < 0.05). In WMLR, BAR was positively associated with kidney stone risk in the unadjusted model (OR = 1.15, 95% CI 1.11–1.18) and after adjustment for demographic, lifestyle, dietary, comorbidity and routine laboratory variables (Model 3; OR = 1.05, 95% CI 1.01–1.10, P = 0.012). After further adjustment for body mass index (BMI), diabetes, estimated glomerular filtration rate (eGFR) and poverty income ratio (PIR) in the fully adjusted primary model (Model 4), the association was attenuated to the null (OR = 0.99, 95% CI 0.95–1.04, P = 0.79); restricting the analysis to participants with eGFR ≥ 60 mL/min/1.73 mZ gave consistent results (OR = 1.01, 95% CI 0.95–1.07, P = 0.78). The apparent nonlinear association observed in the unadjusted analysis (inflection point at BAR = 5.4) was not retained after adjustment (P for nonlinearity = 0.87), and subgroup analyses showed no significant effect modification (all P for interaction > 0.05). In XGBoost, BAR ranked seventh of 28 predictors by gain, with modest discrimination (out-of-fold AUC = 0.61). In the exploratory MIMIC-IV analysis, BAR was positively associated with kidney stones (Model 3 of that cohort: OR = 1.029, 95% CI 1.015–1.045), although this cohort was not designed as a validation set.

**Conclusions:** BAR is not independently associated with kidney stones once renal function and metabolic factors are accounted for, and the previously reported association appears to be largely explained by confounding.

## 1. Introduction

Kidney stones are a prevalent urinary system disease, typically arising from the precipitation of calcium oxalate on the urothelium driven by urine supersaturation, and are clinically characterized by sudden abdominal pain and colic [1]. Despite being standard approaches, treatments such as extracorporeal shock wave lithotripsy and flexible ureteroscopy (FURS) are not without limitations; their effectiveness diminishes when confronting large stones, a scenario often accompanied by prolonged operative duration [2]. The formation of kidney stones involves complex metabolic changes, with dietary factors such as decreased fluid consumption, protein intake, carbohydrates, and oxalate playing key roles [3, 4]. At present, the prevalence of kidney stones continues to rise with a high recurrence rate, leading to decreased quality of life and a substantial healthcare burden [5]. Therefore, it is of vital significance to further identify associate factors for kidney stones and identify at-risk populations to optimize disease management.

The BAR not only serves as a marker of systemic inflammation—where an elevated ratio correlates with worse clinical outcomes—but also provides insight into a patient’s nutritional status and kidney function [6, 7]. Both blood urea nitrogen (BUN) and albumin are intrinsically linked to kidney function, with albumin being abundant in the organic matrix of kidney stones and potentially playing a dual role in calcium oxalate crystallization [8]. In kidney stones patients, elevated BUN is associated with hematuria [9], and albumin(ALB) is negatively correlated with kidney stones prevalence after multivariate adjustment [10], further suggesting a potential association between BAR and kidney stones.

To analyze the association between BAR and kidney stones risk, this study leveraged data from the 2007–2020 National Health and Nutrition Examination Survey (NHANES). Using weighted multivariable logistic regression models and restricted cubic splines, we examined the association between BAR and kidney stones risk, as well as its potential nonlinear relationship. Subsequently, subgroup analyses were conducted to assess the stability of this association and identify relevant at-risk populations. Finally, machine learning methods were utilized to further identify key associate factors for kidney stones, thereby providing a theoretical reference for kidney stones management.

## 2. Materials and methods

### 2.1 Data source

The data of 66,148 participants from 2007 to 2020 were downloaded from the National Health and Nutrition Examination Survey database (NHANES, https://wwwn.cdc.gov/nchs/nhanes/), including outcome variable (kidney stones or not), exposure variable (BAR) and other covariates. Kidney stones-related data were screened based on a questionnaire code [KIQ026: Ever had kidney stones?]. Participants were excluded if they were younger than 18 years or had missing information on the outcome, the exposure or any covariate, including the four additional covariates introduced in this revision (BMI, diabetes, eGFR and PIR). The final analytic sample comprised 23,652 participants, including 2,318 cases with kidney stones and 21,334 controls. More details were shown in Table S1.https://wwwn.cdc.gov/nchs/nhanes/)

### 2.2 Covariates

Specifically, there were six categories of covariates including basic demographic characteristics: age, gender, race, education, marital status and poverty income ratio (PIR); lifestyle and physical activity factors: smoking, drinking, vigorous work activity and activity work moderate; dietary intake factors: intake of sodium, calcium, vitamin A and water; comorbidities: asthma, coronary heart disease, stroke, heart attack and diabetes; anthropometric and renal function measures: body mass index (BMI; kg/mZ) and estimated glomerular filtration rate (eGFR; mL/min/1.73 mZ); and basic hematological and biochemical indicators: mean corpuscular volume (MCV; fL), red blood cell count (RBC; ×10⁶/μL), white blood cell count (WBC; ×103/μL), hemoglobin (g/dL), platelet count (×103/μL), lymphocyte percentage (%), neutrophil percentage (%), aspartate aminotransferase (AST; U/L), alkaline phosphatase (ALP; U/L), and total cholesterol (TC; mg/dL). Diabetes was defined by a self-reported physician diagnosis (DIQ010). eGFR was calculated from serum creatinine using the race-free CKD-EPI 2021 equation [39]. More details were shown in Table S2.

### 2.3 Baseline statistics

To compare baseline characteristics between participants with kidney stones and control participants, categorical variables were described as frequencies (%). Continuous variables following a Gaussian distribution were described as mean ± standard deviation (SD); otherwise, they were described as median (interquartile range, IQR). The gtsummary package (v. 2.4.0) [11] was employed to conduct the baseline analysis and the significant differences were defined by P value < 0.05. The differences between groups were analyzed by Chi-square test and Independent-samples t-test.

### 2.4 Weighted Multivariate Logistic Regression (WMLR) model

To evaluate the independent association between BAR and kidney stones after accounting for potential confounders, we employed weighted multivariable logistic regression models using the survey package (v. 4.4.8) [12]. Four progressively adjusted models were constructed. Model 1 (unadjusted model) included only BAR as the independent variable. Model 2 (partially adjusted model) additionally adjusted for age, sex, and race/ethnicity. Model 3 (conventionally adjusted model) further adjusted for marital status, education level, smoking status, alcohol consumption, dietary intake, and a series of chronic disease histories and laboratory parameters, i.e., the adjustment set used in previous analyses of BAR and kidney stones. Model 4 (fully adjusted, primary model) additionally adjusted for BMI, diabetes, eGFR and PIR. These four variables were included a priori because BUN and albumin are both strongly determined by glomerular filtration and nutritional status, so that a BAR–stone association observed without them cannot be attributed to BAR itself. Model 3 is reported alongside Model 4 to show how the estimate changes once renal function and metabolic status are accounted for, and to allow direct comparison with earlier studies. All models specified a binomial distribution with a logit link function, and hypothesis tests for regression coefficients were based on Wald tests with robust standard errors. Results are presented as odds ratios (ORs) with 95% confidence intervals (CIs). An OR > 1 was considered indicative of a risk factor, an OR < 1 of a protective factor, and two-sided P < 0.05 was considered statistically significant.

### 2.5 Restricted cubic splines (RCS) analysis

To characterize the dose–response relationship between kidney stones and BAR, restricted cubic splines (RCS) with four knots placed at the 5th, 35th, 65th and 95th percentiles of BAR were fitted within the survey-weighted design, and departure from linearity was assessed by a Wald test of the nonlinear spline terms. RCS models were fitted without adjustment and with adjustment for the Model 3 covariates. When nonlinearity was detected, segmented regression was used to estimate the inflection point, i.e., the BAR value at which the slope of the dose–response curve changes. Because a change in slope of a fitted spline does not by itself establish a biological threshold, the inflection point was interpreted descriptively.

### 2.6 Stratification analysis

To further explore the stability of the correlation between kidney stones and BAR in different subgroups, we employed stratification analysis to replay Model 3 in each subgroup. Age was categorized as < 50 or ≥ 50 years, BMI as < 25 or ≥ 25 kg/mZ, PIR as < 1.3 or ≥ 1.3 (the federal poverty threshold), eGFR as < 60 or ≥ 60 mL/min/1.73 mZ, and dietary intakes were dichotomized at their medians. Effect modification was assessed by Wald tests of the BAR × subgroup interaction terms, and a two-sided P for interaction < 0.05 was considered statistically significant. The results were shown using the forestploter package (v. 0.2.0).

### 2.7 Machine learning

To further explore the contribution of different variables in kidney stones, the XGBoost model was employed to analyze each variable in Model 3 using the xgboost package (v. 3.2.1) with learning rate set to 0.5, maximum depth set to 6 and number of iterations set to 25. Variable importance was expressed as Gain, the average improvement in the loss function attributable to a predictor. To avoid optimistic in- sample estimates, discrimination was evaluated using five-fold cross-validated out-of- fold predictions, and the receiver operating characteristic (ROC) curve was plotted using the pROC package (v. 1.18.5) with the area under the curve (AUC). These analyses were exploratory and were not used for inference.

### 2.8 Exploratory analysis in the MIMIC-IV ICU cohort

This analysis was exploratory and hypothesis-generating; it was not designed as, and should not be interpreted as, formal external validation, because MIMIC-IV represents an ICU population that differs substantially from the NHANES general adult population and because kidney stones were ascertained from administrative diagnosis codes rather than from a standardized interview. We included adult patients (≥ 18 years) with their first intensive care unit (ICU) admission and an ICU stay ≥ 24 hours. BAR was calculated from the first available blood urea nitrogen and albumin measurements within 48 hours of ICU admission. Kidney stones were identified by ICD-9 (5920, 5921, 5929) or ICD-10 (N200, N201, N202, N209) diagnosis codes indicating upper urinary tract stones; patients with end-stage renal disease or kidney transplant history were excluded. The same statistical procedures as in the NHANES analysis were applied: weighted multivariate logistic regression (three models), restricted cubic splines with 4 knots, stratification by age and sex, and XGBoost machine learning (learning rate = 0.5, max depth = 6, iterations = 25). All analyses were conducted in R (v. 4.2.2) using the same packages. A total of 20,606 patients (159 with kidney stones, 20,447 controls) from MIMIC-IV were included (Table S3).

### 2.9 Statistical analysis

All statistical analyses were performed using R software (version 4.2.2) with the survey package (v. 4.4.8) to account for the complex sampling design of NHANES; strata containing a single primary sampling unit were handled with the lonely.psu = "adjust" option. Baseline characteristics were compared between participants with and without kidney stones. An OR > 1 was considered indicative of a risk factor, an OR < 1 of a protective factor, and two-sided P < 0.05 was considered statistically significant. In a prespecified sensitivity analysis, participants with eGFR < 60 mL/min/1.73 mZ were excluded, so that moderate-to-severe impairment of renal function could not influence either BAR or the ascertainment of stone disease, and Models 3 and 4 were refitted in the remaining participants. A further set of incremental models added BMI, diabetes, PIR and eGFR to Model 3 one at a time to identify which covariates drove any change in the estimate.

## 3. Results

### 3.1. Baseline features of participants

There were 2,318 participants in the kidney stones group and 21,334 participants in the control group, and the two groups significantly differed in 21 baseline characteristics (P < 0.05) (Table 1). Specifically, compared with the control group, participants with kidney stones were older (53 vs 47 years), and male (55%), non- Hispanic White (77%), married (65%) and people having a smoking habit (51%) or coronary heart disease (5.6%) accounted for larger proportions in the kidney stones group. Participants with kidney stones also had a higher BAR (3.60 vs 3.29), a higher BMI (31 vs 29 kg/mZ), a higher prevalence of diabetes (19% vs 9.3%) and a lower eGFR (91 vs 97 mL/min/1.73 mZ), whereas PIR did not differ between groups (3.09 vs 3.05, P = 0.7). These results initially revealed the potential associate factors of kidney stones and confirmed that renal function and metabolic status differed systematically between the two groups.

**Table 1.** Baseline features of participants.

| **Characteristic** | **Overall (N = 23652)** | **No (N = 21334)** | **Yes (N = 2318)** | **p-value** |
|---|---|---|---|---|
| BAR | 3.32 (1.37) | 3.29 (1.33) | 3.60 (1.66) | <0.001 |
| Age (years) | 48 (17) | 47 (17) | 53 (15) | <0.001 |
| **Gender** |  |  |  | 0.001 |
| Male | 11,664 (49%) | 10,386 (48%) | 1,278 (55%) |  |
| Female | 11,988 (51%) | 10,948 (52%) | 1,040 (45%) |  |
| **Race/ethnicity** |  |  |  | <0.001 |
| Mexican American | 3,317 (8.0%) | 3,034 (8.2%) | 283 (6.2%) |  |
| Other Hispanic | 2,341 (5.8%) | 2,080 (5.8%) | 261 (5.6%) |  |
| Non-Hispanic White | 10,181 (69%) | 8,903 (68%) | 1,278 (77%) |  |
| Non-Hispanic Black | 4,997 (10%) | 4,694 (11%) | 303 (5.4%) |  |
| Other Race | 2,816 (7.6%) | 2,623 (7.8%) | 193 (6.1%) |  |
| **Education** |  |  |  | 0.310 |
| Less than high school | 4,952 (13%) | 4,442 (13%) | 510 (14%) |  |
| High school or above | 18,700 (87%) | 16,892 (87%) | 1,808 (86%) |  |
| **Marital status** |  |  |  | <0.001 |
| Married | 12,625 (58%) | 11,272 (57%) | 1,353 (65%) |  |
| Other | 6,301 (24%) | 5,611 (24%) | 690 (24%) |  |
| Never married | 3,276 (12%) | 3,115 (13%) | 161 (6.4%) |  |
| Living with partner | 1,450 (5.6%) | 1,336 (5.7%) | 114 (4.7%) |  |
| Vigorous work activity | 5,024 (23%) | 4,483 (23%) | 541 (26%) | 0.023 |
| Moderate work activity | 9,310 (44%) | 8,410 (44%) | 900 (43%) | 0.538 |
| Sodium intake (mg) | 3,577 (1,794) | 3,579 (1,780) | 3,560 (1,912) | 0.213 |
| Calcium intake (mg) | 980 (589) | 984 (593) | 945 (557) | 0.046 |
| Vitamin A intake (mcg) | 638 (702) | 637 (694) | 647 (769) | 0.933 |
| Water intake (g) | 3,096 (1,551) | 3,100 (1,557) | 3,056 (1,490) | 0.426 |
| Smoking | 10,444 (44%) | 9,263 (43%) | 1,181 (51%) | <0.001 |
| Alcohol drinking | 18,340 (83%) | 16,548 (83%) | 1,792 (84%) | 0.595 |
| Asthma | 3,599 (15%) | 3,167 (15%) | 432 (18%) | 0.009 |
| Coronary heart disease | 983 (3.7%) | 799 (3.4%) | 184 (5.6%) | <0.001 |
| Stroke | 903 (2.9%) | 774 (2.8%) | 129 (3.9%) | 0.009 |
| Heart attack | 985 (3.3%) | 800 (3.0%) | 185 (5.5%) | <0.001 |
| BMI (kg/m²) | 29 (7) | 29 (7) | 31 (7) | <0.001 |
| Diabetes | 3,149 (10%) | 2,623 (9.3%) | 526 (19%) | <0.001 |
| eGFR (mL/min/1.73 m²) | 96 (21) | 97 (21) | 91 (21) | <0.001 |
| PIR | 3.06 (1.65) | 3.05 (1.65) | 3.09 (1.61) | 0.654 |
| MCV (fL) | 89.5 (5.4) | 89.5 (5.4) | 89.6 (5.4) | 0.163 |
| RBC (million cells/µL) | 4.69 (0.48) | 4.69 (0.48) | 4.72 (0.46) | 0.006 |
| WBC (1000 cells/µL) | 7.26 (3.15) | 7.24 (3.22) | 7.45 (2.39) | 0.009 |
| Hemoglobin (g/dL) | 14.21 (1.45) | 14.19 (1.45) | 14.30 (1.43) | 0.007 |
| Platelet (1000 cells/µL) | 240 (61) | 241 (60) | 239 (64) | 0.088 |
| Lymphocyte percentage (%) | 30 (8) | 30 (8) | 29 (8) | <0.001 |
| Neutrophil percentage (%) | 58 (9) | 58 (9) | 59 (9) | 0.001 |
| AST (U/L) | 25 (15) | 25 (16) | 25 (12) | 0.438 |
| ALP (U/L) | 69 (24) | 69 (24) | 72 (22) | <0.001 |
| Total cholesterol (mg/dL) | 192 (41) | 192 (41) | 190 (44) | 0.137 |

### 3.2. Association between BAR and kidney stones risk

The four models revealed a graded attenuation of the association between kidney stones and BAR (Table 2). In the unadjusted model, each one-unit increase in BAR was associated with a 15% higher odds of kidney stones (OR = 1.15, 95% CI 1.11–1.18, P < 0.001). With the conventional adjustment set used in previous studies (Model 3), the association remained statistically significant (OR = 1.05, 95% CI 1.01–1.10, P = 0.012). After additional adjustment for BMI, diabetes, eGFR and PIR in the fully adjusted primary model (Model 4), however, the estimate moved to the null (OR = 0.99, 95% CI 0.95–1.04, P = 0.79). In the sensitivity analysis restricted to participants with eGFR ≥ 60 mL/min/1.73 mZ (n = 21,819), the corresponding estimates were OR = 1.06 (95% CI 0.99–1.13, P = 0.079) in Model 3 and OR = 1.01 (95% CI 0.95–1.07, P = 0.78) in Model In the incremental models, adding BMI alone reduced the Model 3 estimate to OR = 1.03 (95% CI 0.99–1.08, P = 0.13), adding diabetes to OR = 1.04 (95% CI 0.99–1.08, P = 0.063) and adding eGFR to OR = 1.03 (95% CI 0.98–1.07, P = 0.24), whereas adding PIR alone did not materially change it (OR = 1.05, 95% CI 1.01–1.10, P = 0.011) (Table S4). In the unadjusted RCS analysis, BAR was related to kidney stones in a nonlinear manner (P < 0.001), with an inflection point at BAR = 5.4: the odds rose steeply below this value and more gradually above it (Figure 1A). After adjustment for the Model 3 covariates, the nonlinear component was no longer significant (P = 0.87) and the curve was essentially linear and flat (Figure 1B). In summary, BAR was associated with kidney stones in models that did not account for renal function and metabolic status, but this association did not persist in the fully adjusted model.

**Figure 1.**
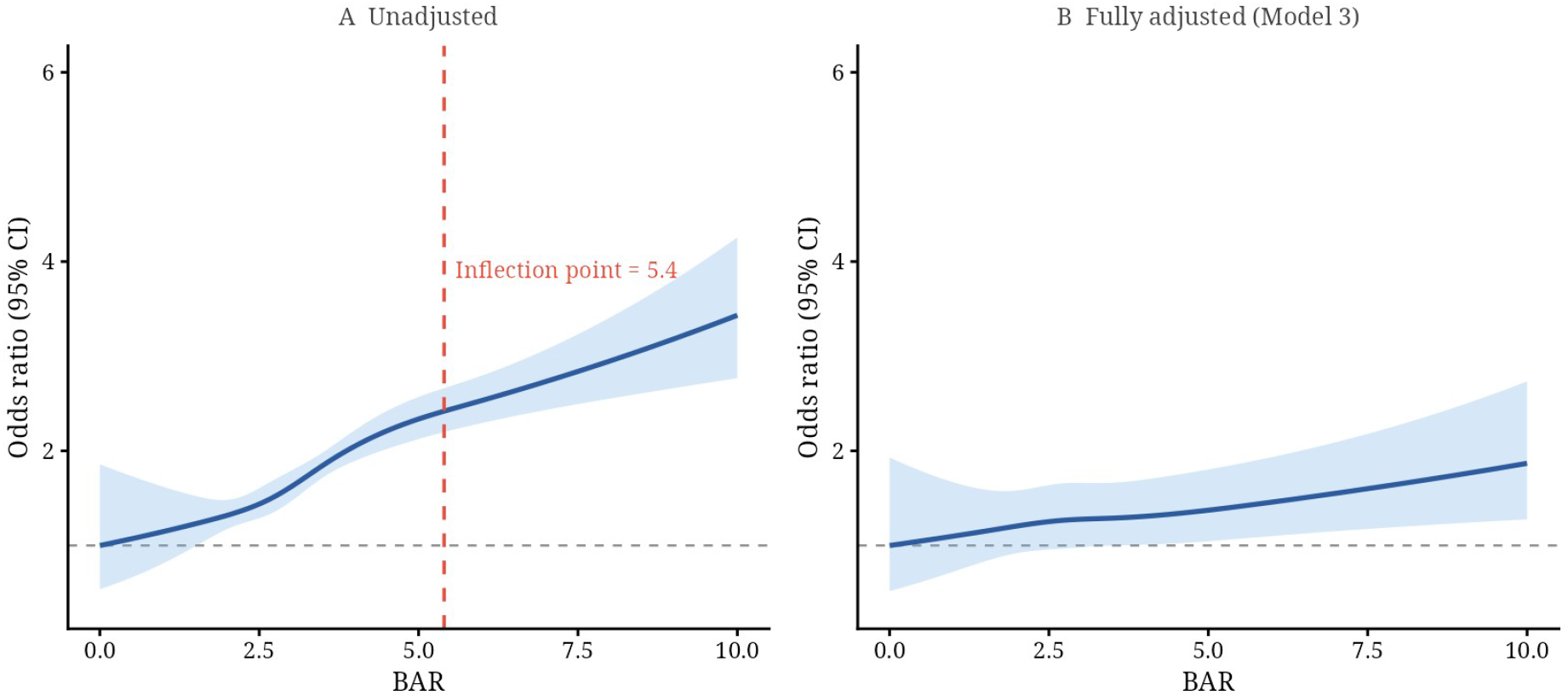
Restricted cubic spline analysis of the association between kidney stones and BAR. (A) Unadjusted model; (B) Model 3, conventionally adjusted for demographic, lifestyle, dietary, comorbidity and routine laboratory variables. The solid line represents the odds ratio (OR) and the shaded area the 95% confidence interval (CI), with the minimum BAR as the reference. Four knots were placed at the 5th, 35th, 65th and 95th percentiles of BAR. A significant nonlinear association was observed in the unadjusted model (P < 0.001), with an inflection point at BAR = 5.4 (dashed red line); after adjustment the nonlinear component was not significant (P = 0.87). The x-axis is truncated at 10 (99.5th percentile) for legibility.

**Table 2.**
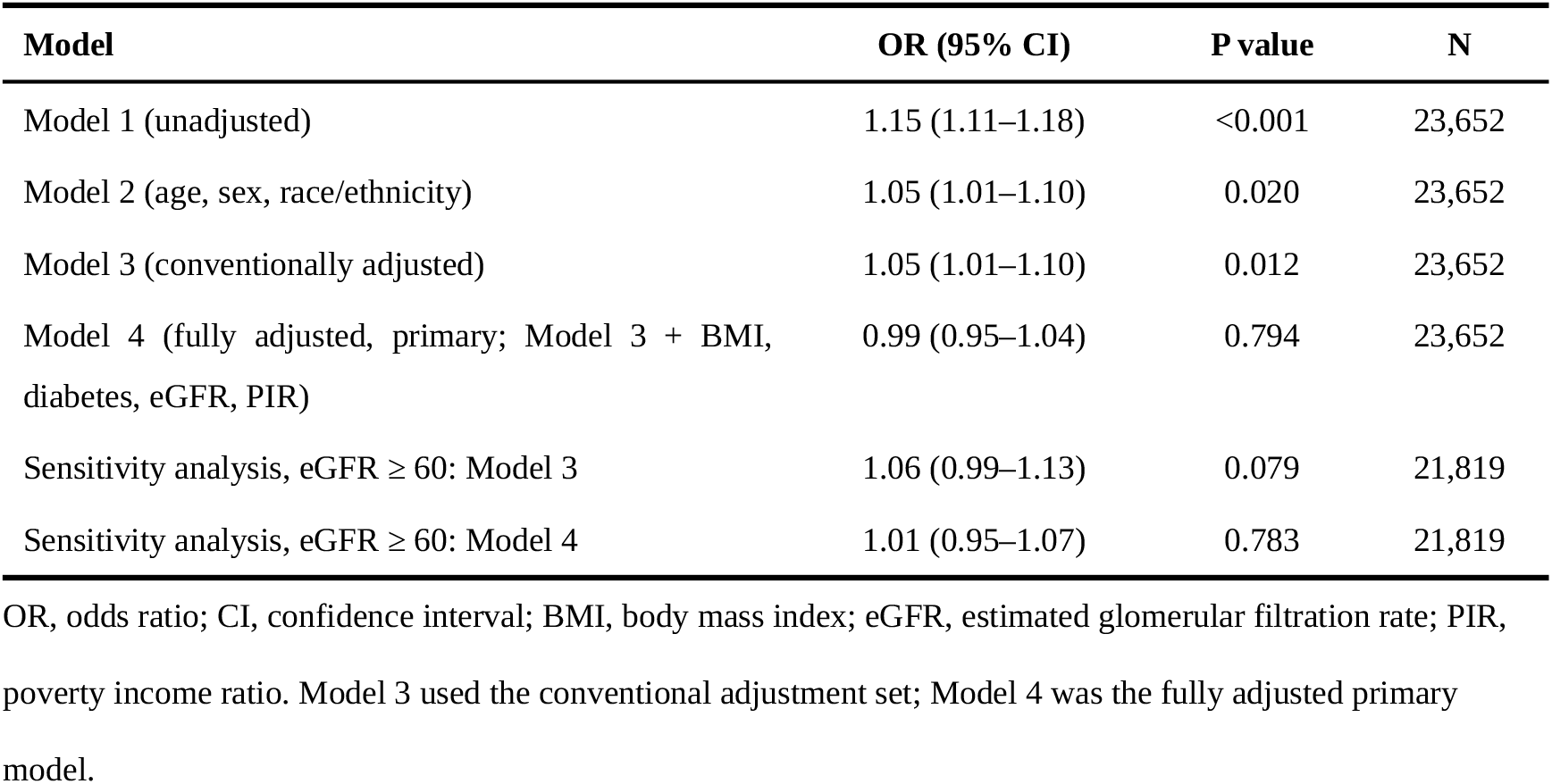
Association between kidney stones and BAR in four models, and in the sensitivity analysis restricted to participants with eGFR ≥ 60 mL/min/1.73 m².

| **Model** | **OR (95% CI)** | **P value** | **N** |
|---|---|---|---|
| Model 1 (unadjusted) | 1.15 (1.11–1.18) | <0.001 | 23,652 |
| Model 2 (age, sex, race/ethnicity) | 1.05 (1.01–1.10) | 0.020 | 23,652 |
| Model 3 (conventionally adjusted) | 1.05 (1.01–1.10) | 0.012 | 23,652 |
| Model 4 (fully adjusted, primary; Model 3 + BMI, diabetes, eGFR, PIR) | 0.99 (0.95–1.04) | 0.794 | 23,652 |
| Sensitivity analysis, eGFR ≥ 60: Model 3 | 1.06 (0.99–1.13) | 0.079 | 21,819 |
| Sensitivity analysis, eGFR ≥ 60: Model 4 | 1.01 (0.95–1.07) | 0.783 | 21,819 |

OR, odds ratio; CI, confidence interval; BMI, body mass index; eGFR, estimated glomerular filtration rate; PIR, poverty income ratio. Model 3 used the conventional adjustment set; Model 4 was the fully adjusted primary model.

### 3.3. Subgroup and machine-learning analyses

In the stratified analyses the direction of the association was consistent across subgroups, but no subgroup showed statistically significant effect modification (all P for interaction > 0.05) (Figure 2A–B). The largest point estimates were observed in females (OR = 1.12, 95% CI 1.06–1.17), participants with less than a high school education (OR = 1.10, 95% CI 1.01–1.20), participants with diabetes (OR = 1.09, 95% CI 1.02–1.17) and those with a PIR < 1.3 (OR = 1.07, 95% CI 1.02–1.13), whereas the estimates in males (OR = 1.01, 95% CI 0.95–1.08) and in participants with eGFR < 60 mL/min/1.73 mZ (OR = 1.05, 95% CI 0.99–1.12) were close to the null. Because none of the interaction terms reached significance, these differences should be read as variation in precision rather than as evidence that any group is specifically susceptible to BAR- related stone risk. In the exploratory XGBoost model built on the Model 3 variables, BAR ranked seventh of 28 predictors by gain (0.057) (Figure 3), behind age, neutrophil percentage, MCV, WBC count, vitamin A intake and RBC count, and discrimination was modest (five-fold out-of-fold AUC = 0.61, 95% CI 0.60–0.63) (Figure 4). These results indicate that BAR carries little predictive information for kidney stones beyond routinely available clinical variables.

**Figure 2.**
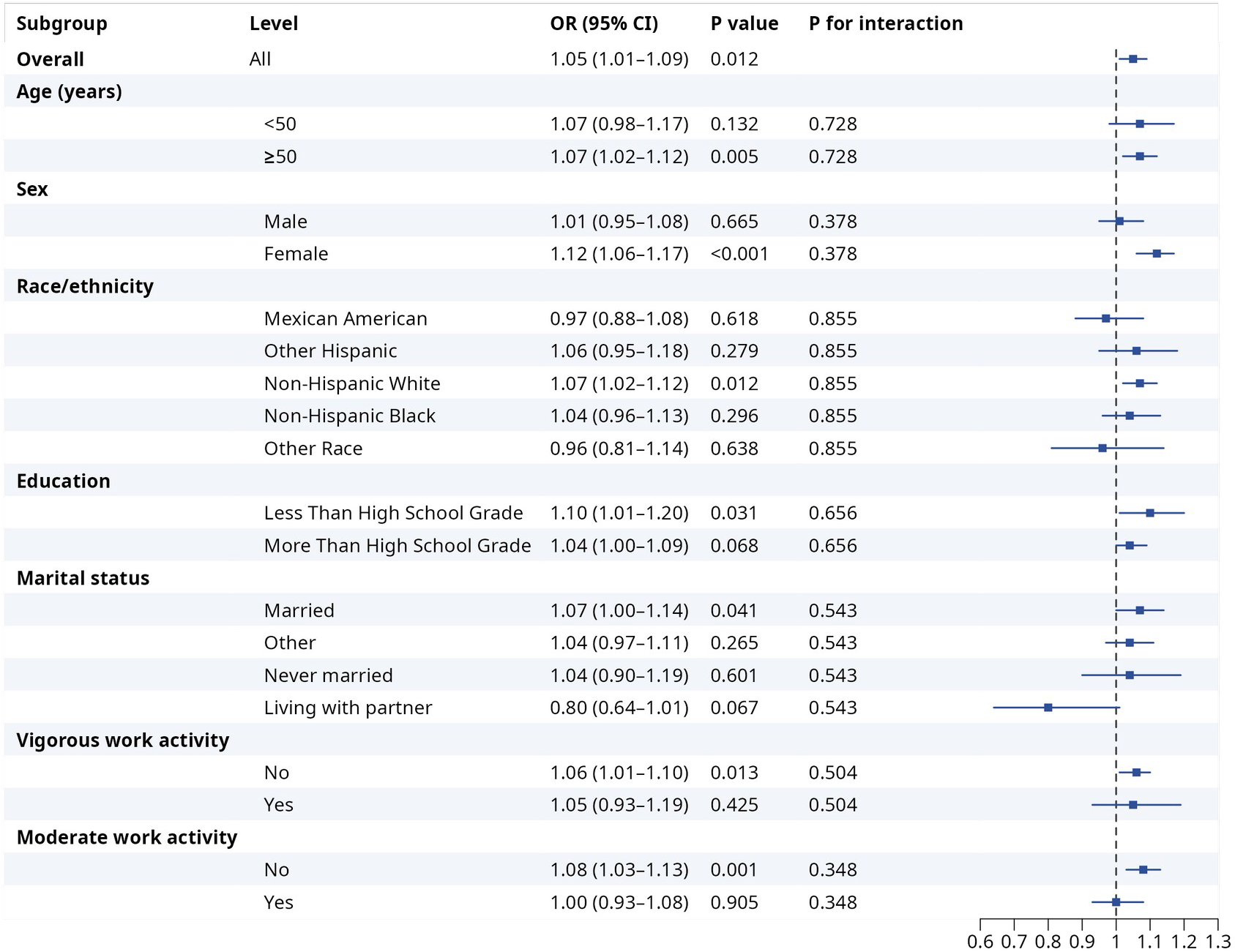
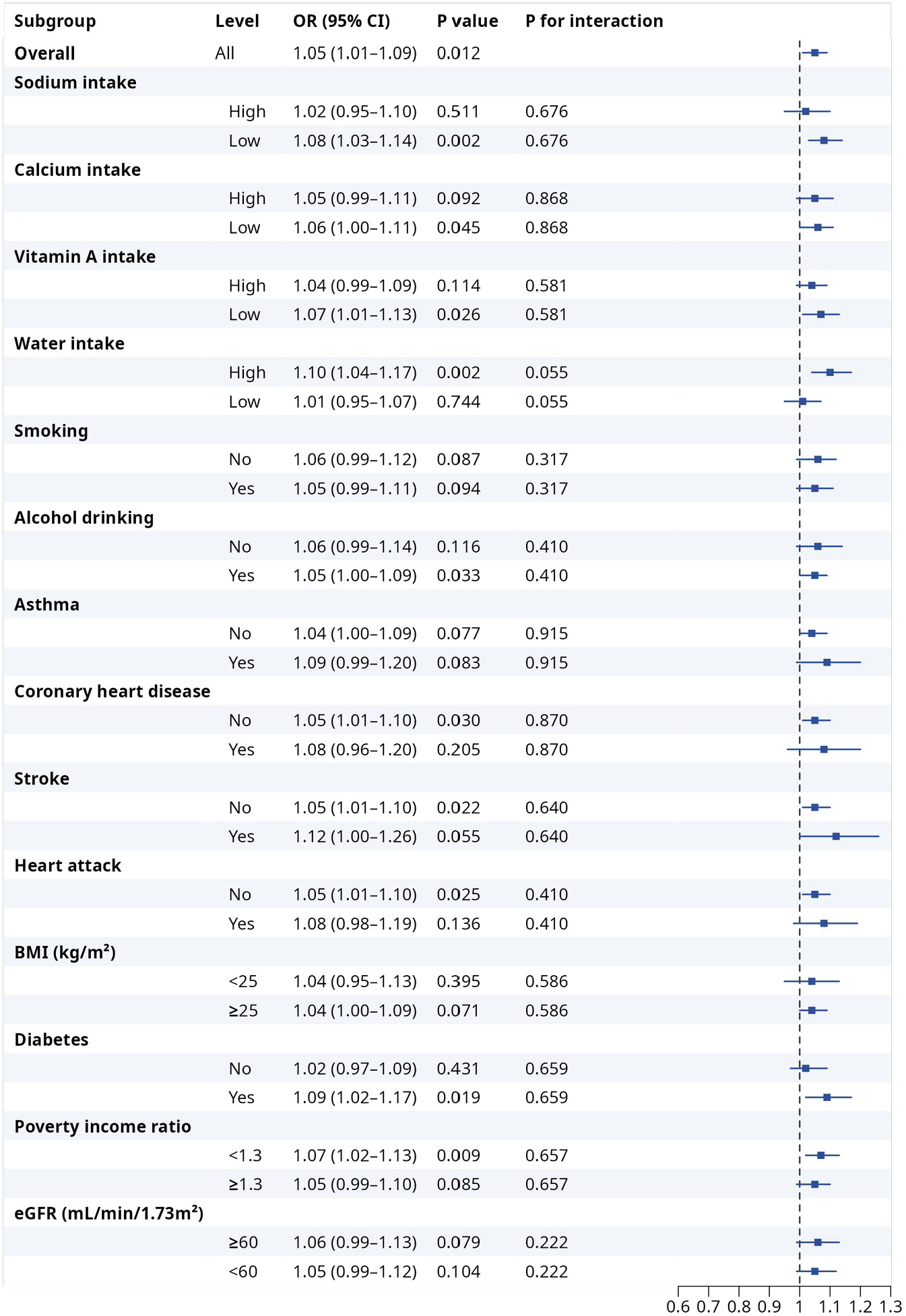
Association between kidney stones and BAR in prespecified subgroups (Model 3). (A) Demographic and physical activity subgroups; (B) dietary, lifestyle, comorbidity and metabolic subgroups. Squares represent odds ratios per one-unit increase in BAR and horizontal lines the 95% confidence intervals; the dashed vertical line indicates OR = 1. P for interaction was obtained from Wald tests of the BAR × subgroup product terms.

**Figure 3.**
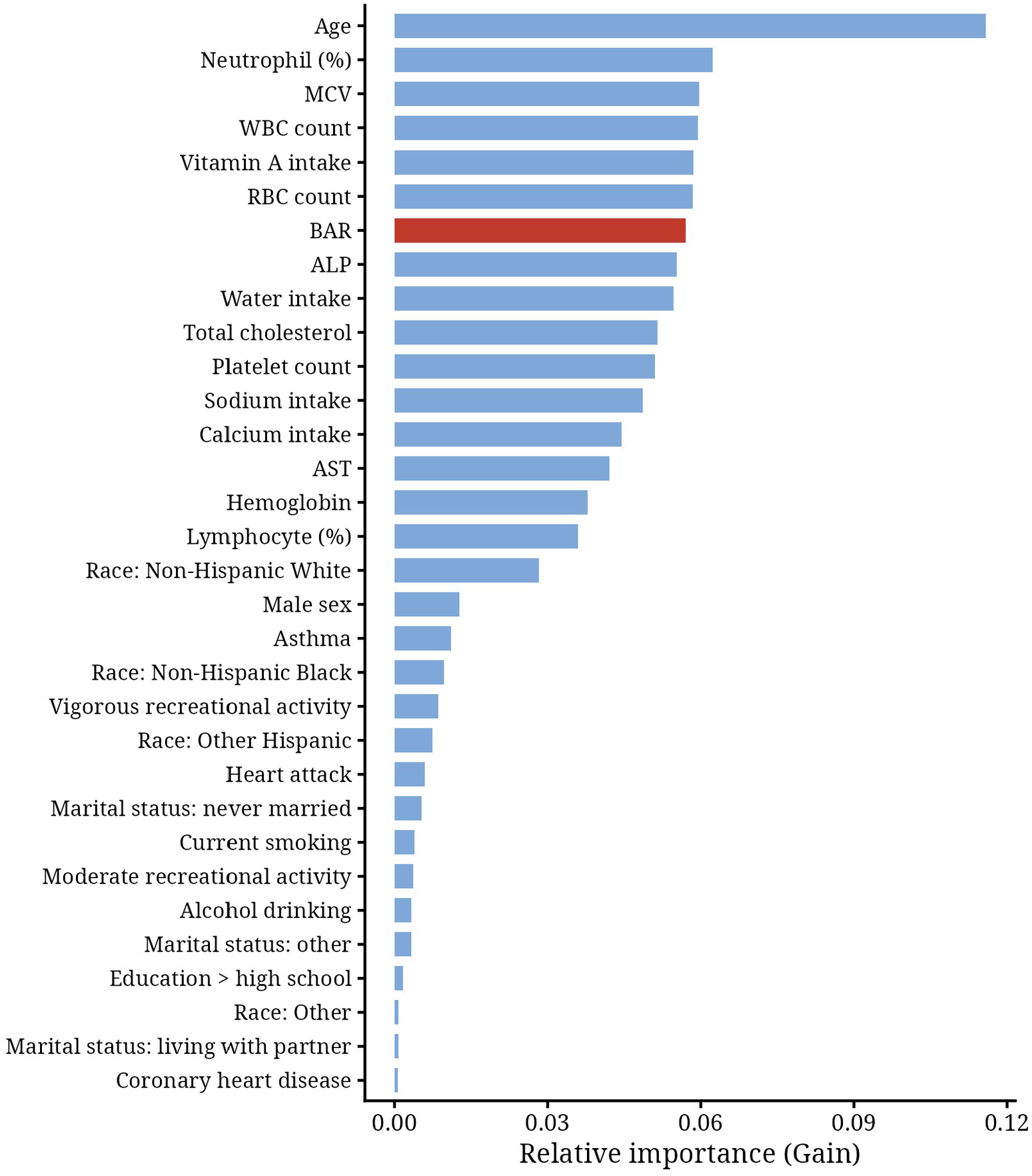
Importance rank of variables in the XGBoost model. Bars show Gain, the average improvement in the loss function attributable to each predictor; BAR is highlighted in red.

**Figure 4.**
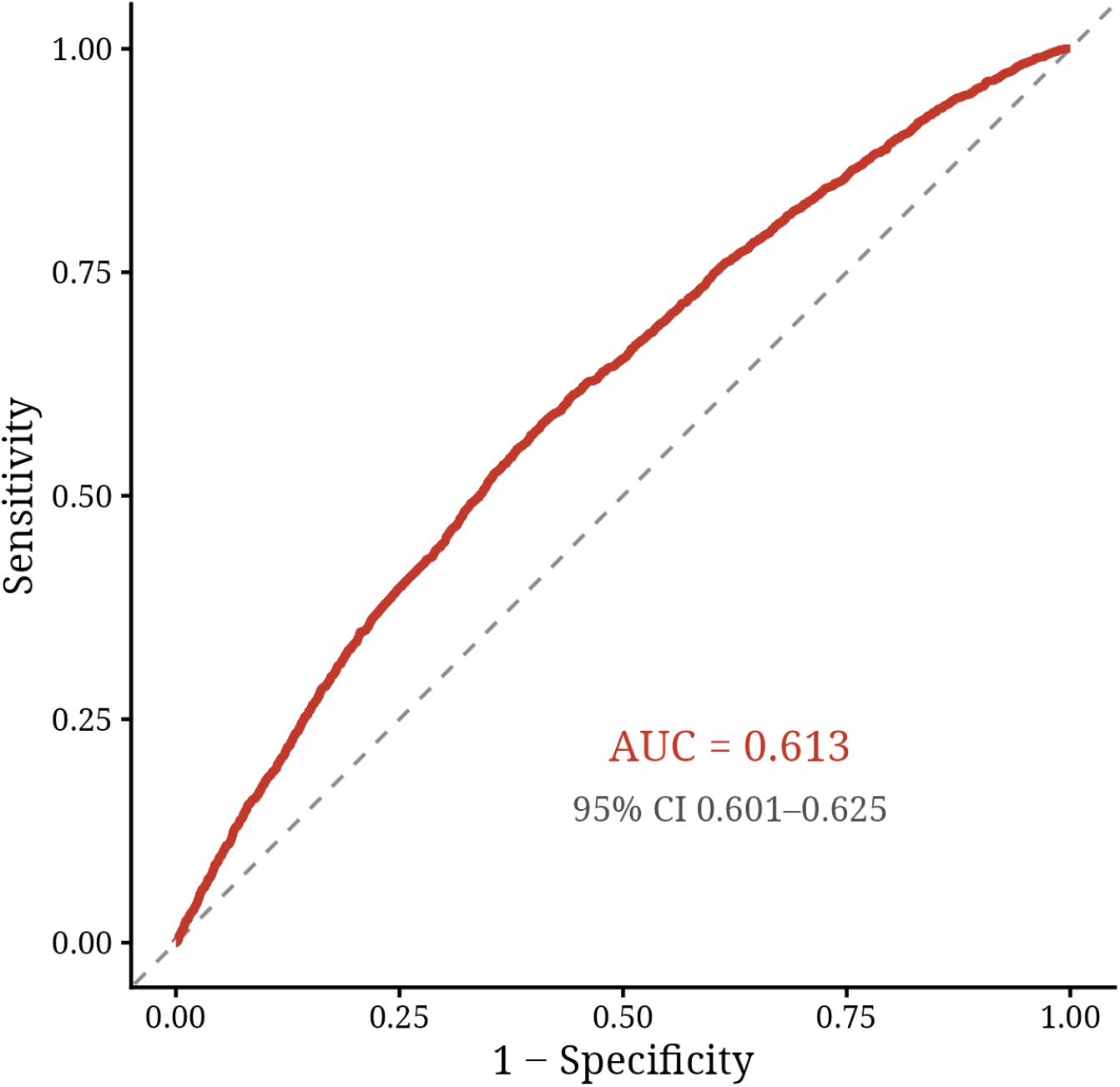
Receiver operating characteristic curve of the XGBoost model, based on five-fold out-of-fold predictions.

### 3.4. Exploratory analysis in the MIMIC-IV ICU cohort

In multivariable logistic regression, BAR was positively associated with kidney stones in all three models of this exploratory analysis, which are numbered independently of the NHANES models (Model 1 OR = 1.029, 95% CI 1.016–1.043, P < 0.001; Model 2 OR = 1.030, 95% CI 1.016–1.044, P < 0.001; Model 3 OR = 1.029, 95% CI 1.015–1.045, P < 0.001) (Table 3). The RCS analysis suggested a nonlinear dose- response relationship (nonlinear P = 0.0006), with an upward trend similar in direction to that seen in the NHANES cohort (Figure 5; Table 4). Subgroup analyses showed that BAR was significantly associated with kidney stones in patients aged ≥ 60 years (OR = 1.031, 95% CI 1.014–1.049, P < 0.001), in males (OR = 1.027, 95% CI 1.008–1.047, P = 0.006) and in females (OR = 1.034, 95% CI 1.010–1.058, P = 0.005), with all subgroup ORs > 1 (Figure 6; Table 5). The XGBoost model identified BAR as the most important predictor of kidney stones (Gain = 0.265), ahead of other clinical covariates (Figure 7; Table 6). Because this cohort was assembled from administrative ICU data with a small number of stone cases, these findings are exploratory: they show that the direction of the association is reproducible in a clinically distinct population, but they neither confirm nor refute the NHANES results.

**Figure 5.**
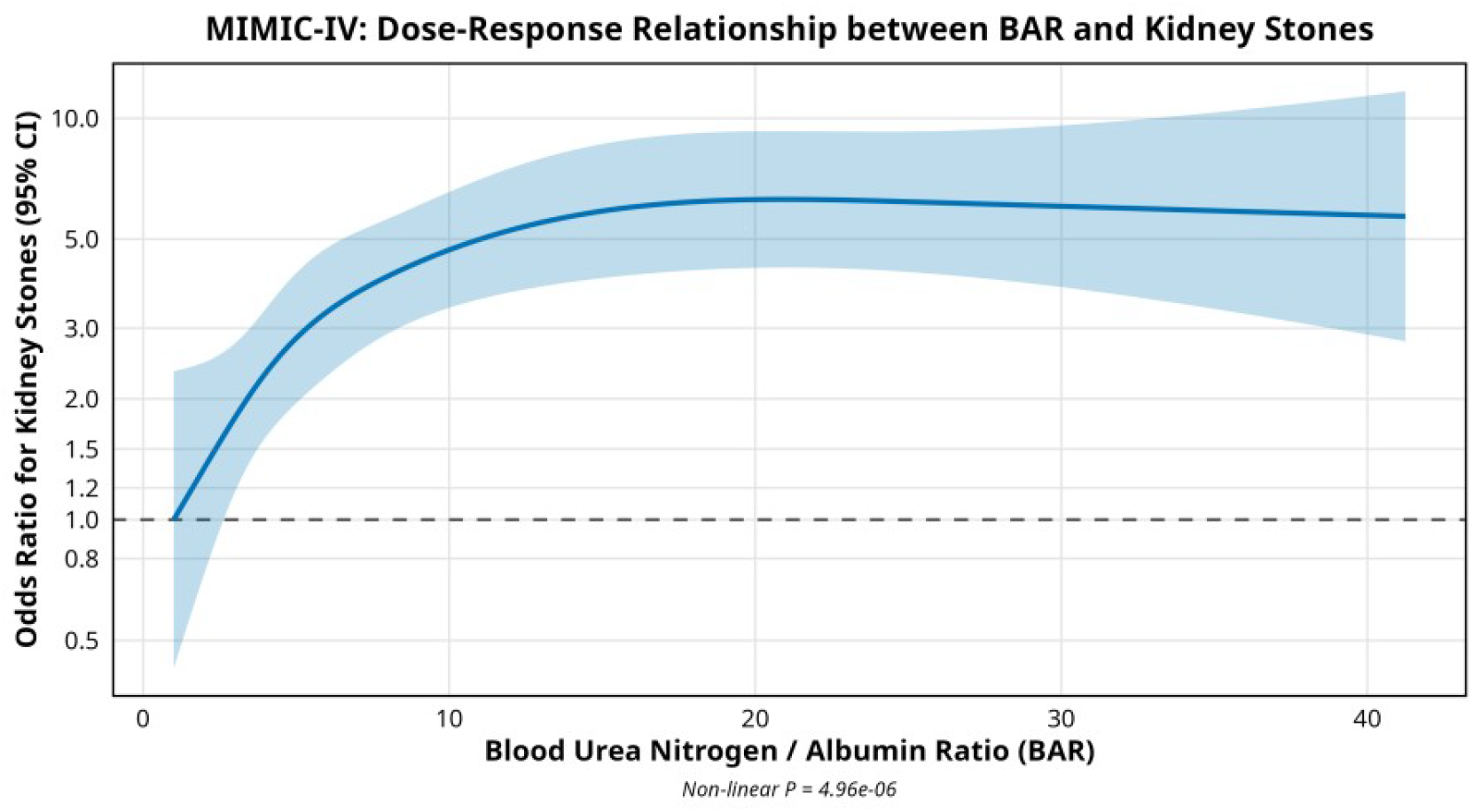
Restricted cubic spline analysis of the association between kidney stones and BAR in the MIMIC-IV exploratory cohort.

**Figure 6.**
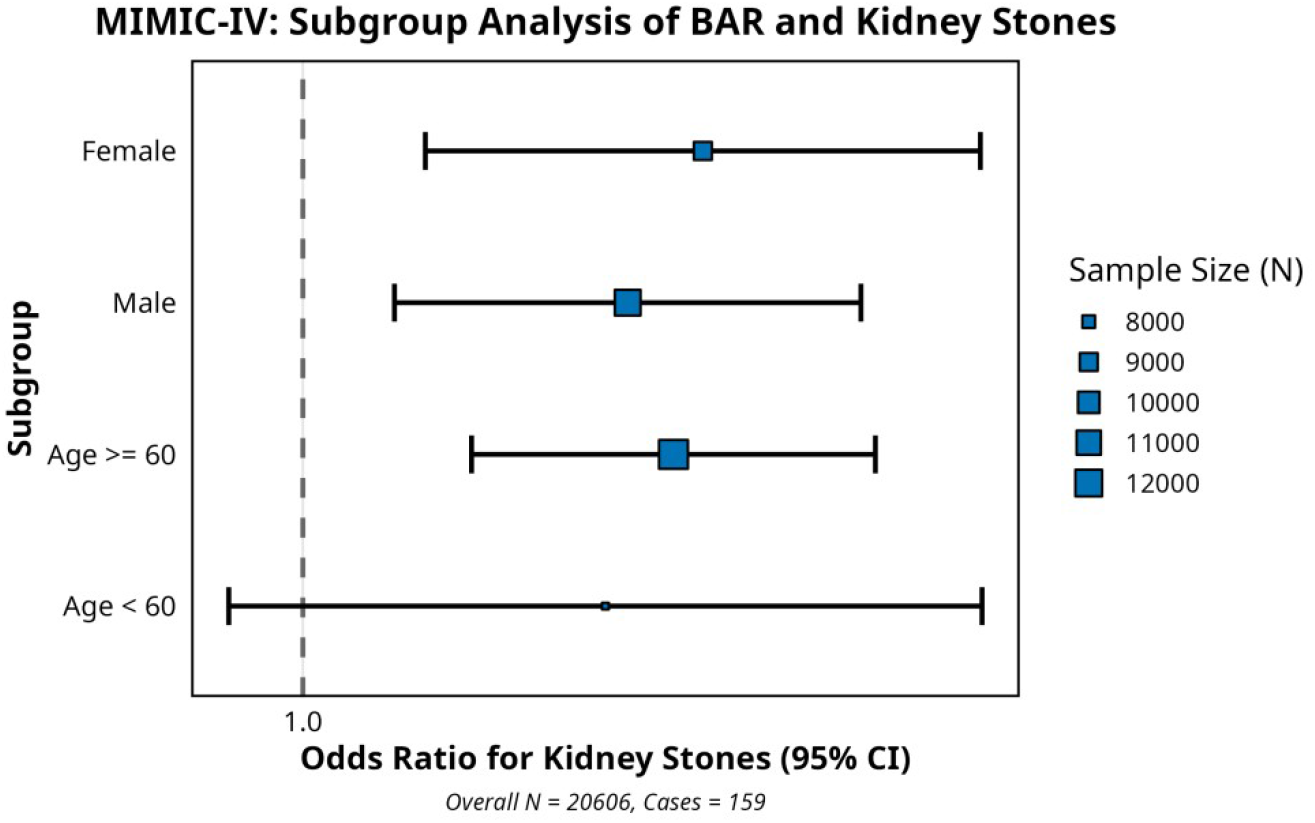
**Association between kidney stones and BAR in subgroups of the MIMIC-IV exploratory cohort.**

**Figure 7.**
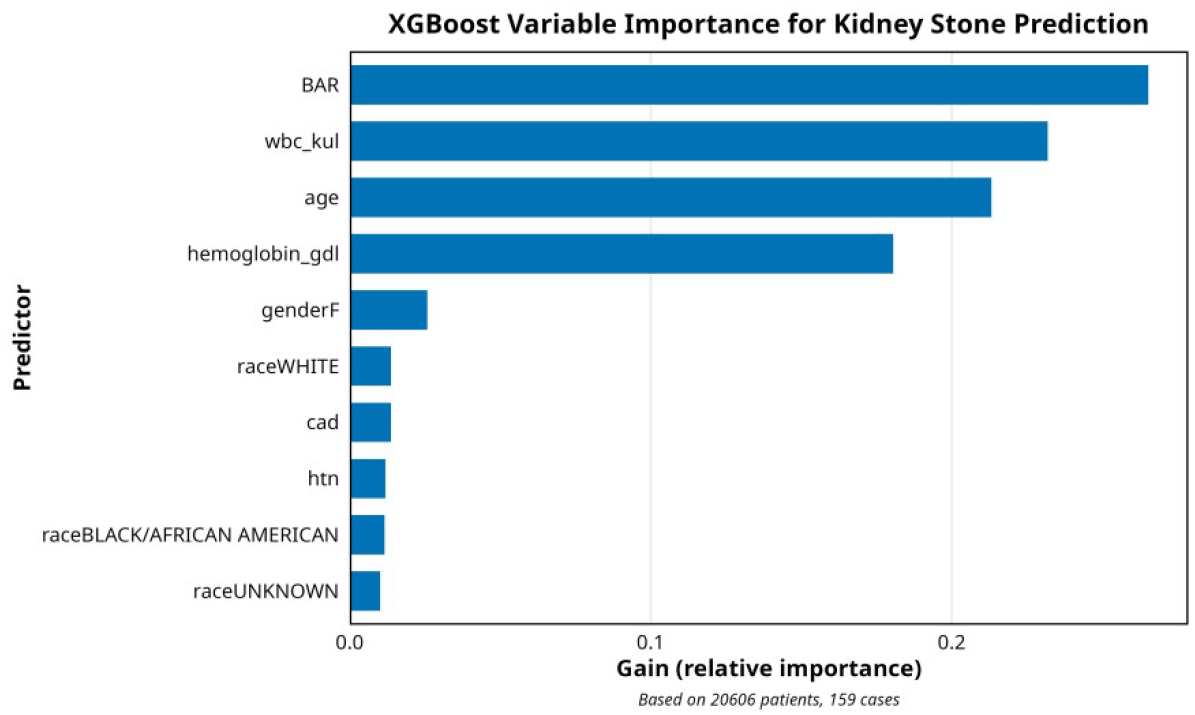
Importance rank of variables in the XGBoost model, MIMIC-IV exploratory cohort.

**Table 3.** Association between kidney stones and BAR in the MIMIC-IV exploratory cohort.

| Model | OR | 95% CI | P value | N |
|---|---|---|---|---|
| Model 1 | 1.029 | 1.016–1.043 | 2.24×10<sup>−5</sup> | 20,606 |
| Model 2 | 1.03 | 1.016–1.044 | 3.45×10<sup>−5</sup> | 20,606 |
| Model 3 | 1.029 | 1.015–1.045 | 1.03×10<sup>−4</sup> | 20,606 |

**Table 4.**
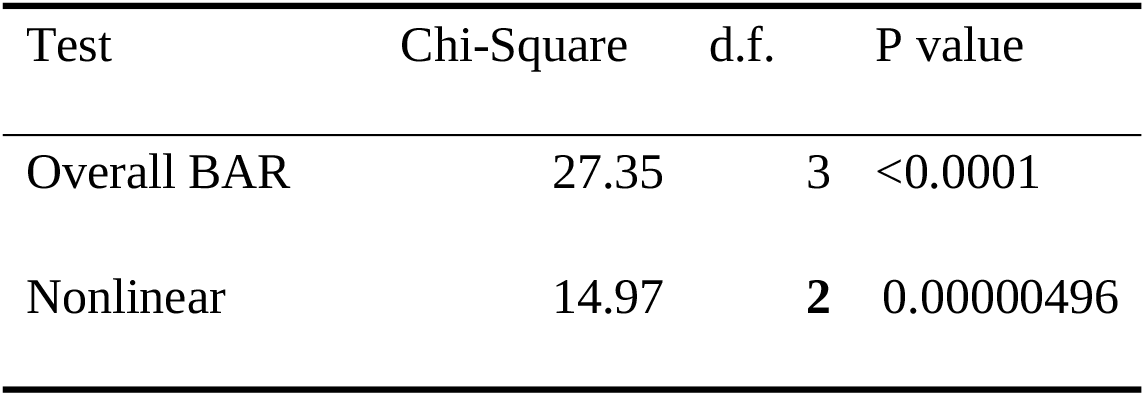
Restricted cubic spline analysis in the MIMIC-IV exploratory cohort.

**Table 5.** Subgroup analysis in the MIMIC-IV exploratory cohort.

| Subgroups | OR | 95% CI | P value | N |
|---|---|---|---|---|
| **Age < 60** | 1.025 | 0.994–1.058 | 0.115 | 7,621 |
| **Age ≥ 60** | 1.031 | 1.014–1.049 | 0.0003 | 12,985 |
| **Male** | 1.027 | 1.008–1.047 | 0.006 | 11,588 |
| **Female** | 1.034 | 1.010–1.058 | 0.005 | 9,018 |

**Table 6.** Gain values of the ranked features in the MIMIC-IV exploratory cohort.

| Rank | Features | Gain |
|---|---|---|
| 1 | BAR | 0.265 |
| 2 | WBC count | 0.232 |
| 3 | Age | 0.213 |
| 4 | Hemoglobin | 0.181 |
| 5 | Female sex | 0.026 |
| 6 | Race: White | 0.014 |
| 7 | Coronary artery disease | 0.014 |
| 8 | Hypertension | 0.012 |
| 9 | Race: Black/African American | 0.012 |
| 10 | Race: unknown | 0.010 |

## 4. Discussion

The BAR is a composite biomarker reflecting inflammatory status, nutritional condition, and renal function [6, 7]. Its components, BUN and albumin, are intrinsically linked to kidney physiology [8, 13]; elevated BUN has been associated with hematuria in patients with kidney stones, and albumin is negatively correlated with kidney stone prevalence after multivariate adjustment [10]. These observations motivated the present analysis, in which kidney stone formers had higher BAR levels and also differed from controls in several conventional risk factors. Weighted multivariable logistic regression showed a positive association in the unadjusted model and after adjustment for a broad set of demographic, lifestyle, dietary, comorbidity and laboratory variables (Model 3), but the association was attenuated to the null once BMI, diabetes, eGFR and PIR were added (Model 4). The apparent nonlinear relationship, and the inflection point at approximately 5.4 seen in the unadjusted analysis, were likewise not retained after adjustment. Taken together, these results suggest that the crude association between BAR and kidney stones largely reflects confounding by renal function and metabolic status, rather than an independent effect of BAR. Age is a well-established risk factor for kidney stones [5, 14]. Kidney stones patients typically have a mean age of 47 ± 15 years [15], and age-related oxidative stress may contribute to this increase in risk [16]. Notably, oxidative stress triggered by calcium oxalate crystals can induce premature senescence of renal tubular epithelial cells, thereby exacerbating crystal deposition [17]. On the other hand, vitamin D deficiency is common in older adults and is associated with impaired bone health [18], while both excessive and insufficient vitamin D intake may affect renal function, highlighting the need for careful supplementation in this population. Sedentary behavior, often associated with limited mobility in older individuals, further contributes to kidney stones risk.

An observed gender disparity exists in kidney stones prevalence, with older males exhibiting higher rates than premenopausal women. This disparity may stem from hormonal and metabolic differences, particularly the role of estrogen in downregulating receptors for calcium oxalate crystal adhesion [19, 20]. Thus, hormonal modulation may offer protective effects in females, while males require more targeted preventive strategies.

Neutrophils are key infiltrators in inflamed tissues. They play a central role in pathogen clearance through mechanisms such as neutrophil extracellular trap (NET) release and fatty acid metabolism-dependent reactive oxygen species (ROS) generation. However, they have been associated with the induction of chronic inflammation and the aggravation of tissue injury [21, 22]. In the context of kidney stones, metabolic disorders and inflammation are critical drivers, with neutrophils potentially mediating the effects of lipid aggregates on stone formation [23]. Moreover, neutrophil gelatinase- associated lipocalin (NGAL) secreted by neutrophils promotes calcium oxalate crystallization, crystal growth, aggregation, adhesion to renal cells, and extracellular matrix invasion. Therefore, neutrophils may serve as key indicators of inflammatory pathology in kidney stones, and our observation that neutrophil percentage ranked second by gain in the XGBoost model is consistent with this mechanistic evidence.

Smoking, including secondhand exposure, is established as a significant behavioral risk factor contributing to the development of kidney stones [3, 24]. Cigarette smoke contains over 7,000 chemicals, including oxidizing gases and heavy metals, which induce oxidative stress and sustain neutrophil recruitment [25, 26], thereby exacerbating kidney stones-related inflammation [1, 27].

Furthermore, an elevated risk of cardiovascular diseases—such as stroke, hypertension, and CHD—has been observed in patients with kidney stones. This association may involve mechanisms such as vascular calcification, osteopontin- induced oxidative stress, and endothelial dysfunction [28, 29].

These mechanisms concern stone formation in general rather than the incremental value of BAR itself. In our data, the association did not differ materially across subgroups defined by sex, smoking status, cardiovascular disease or age (all P for interaction > 0.05), and adjusting for these variables in Model 3 did not attenuate the estimate; the attenuation appeared only when renal function and metabolic status were accounted for.

The attenuation observed after adjustment for eGFR is biologically plausible. BAR is the ratio of BUN to albumin, and both analytes are strongly determined by glomerular filtration and by the patient’s nutritional and inflammatory state [6, 7]. BUN rises with reduced renal perfusion and impaired filtration, so BAR partly encodes renal function; eGFR is therefore not a neutral covariate but one that shares a common cause with the exposure. When eGFR, BMI and diabetes were added, the BAR estimate moved towards unity, which is the pattern expected when an observed association is confounded by these factors rather than generated by the exposure itself. The incremental models showed that BMI, diabetes and eGFR each contributed to this attenuation, whereas PIR did not. BAR therefore retains descriptive value as an index that combines nutritional and inflammatory information, but our data do not support its use as an independent risk marker for stones once renal function and metabolic status are known.

Younger individuals represent a growing at-risk population for kidney disease, a trend driven in part by dietary shifts. This age group also carries a substantial burden of hereditary kidney stones, which often present at a mean age of 30 ± 14 years [15], further underscoring the early onset of stone-related pathology. In addition to genetic predisposition, younger populations exhibit a higher prevalence of nutritional risk and elevated blood urea nitrogen-to-creatinine ratios despite lower rates of normal nutritional status [30]. Moreover, adolescents and young adults show heightened susceptibility to the renal effects of long-term inorganic pollutant exposure, which has been correlated with BUN and albumin-to-creatinine ratios [31]. Collectively, these findings suggest that multiple pathways—dietary, genetic, nutritional, and environmental—converge to elevate kidney stone risk in young populations. Previous analyses have suggested that indices combining BUN with nutritional markers may be particularly informative in young, malnourished populations [32]. In our data, however, the BAR–stone estimate did not differ between participants younger and older than 50 years (P for interaction = 0.73; OR 1.07 in both groups), so we found no evidence of age-specific usefulness. The rising burden of stone disease in this group is better addressed through the dietary and metabolic pathways described above.

Although the point estimate was higher in females than in males, the interaction test was not significant (P = 0.38), and the difference is more plausibly explained by differences in case numbers and precision than by a true sex-specific effect. The same caution applies to the other subgroups in which the estimate happened to reach significance. Previous reports linking BAR to mortality after surgery in critically ill patients [33] concerned a different outcome and a different population, and our data do not provide support for effect modification by sex in the stone setting.

The role of fluid intake in the development of kidney stones is complex and highly dependent on individual metabolic status and dietary habits [4]. Although inadequate hydration is a classic risk factor, the type of beverage consumed modulates this effect: magnesium-rich water may reduce stone risk, whereas hard water with high calcium content has been associated with increased risk in women over 60 [34]. Consumption of sugar-sweetened beverages, including apple juice, induces elevations in insulin, BUN, and lactate levels, and although increased water intake generally lowers BUN [35], higher bottled water consumption has been paradoxically associated with increased kidney stones prevalence [36, 37]. In patients with pneumonia, excessive fluid administration may lead to overload because of impaired water excretion [38]. In our data the point estimate was higher in participants with high water intake (OR = 1.10, 95% CI 1.04–1.17) than in those with low intake (OR = 1.01, 95% CI 0.95–1.07), but the interaction term did not reach significance (P = 0.055), so this pattern should be regarded as hypothesis-generating. Attention to water quality, with preference given to soft water or beverages with higher magnesium content, remains reasonable advice.

However, several limitations should be noted. First, the cross-sectional nature of the NHANES dataset prevents causal inferences regarding the relationship between BAR and kidney stones, and reverse causation cannot be excluded. Second, the use of self-reported data for stone history and for several other variables may have introduced recall and misclassification bias, and stone episodes are likely to have been under-ascertained. Third, BAR was derived from a single measurement of BUN and albumin and therefore carries within-person variability. Fourth, although eGFR was included as a covariate, a single creatinine-based estimate cannot fully capture renal tubular function or the prerenal components of BUN, so residual confounding by renal function remains possible; conversely, because BUN is itself a filtration marker, adjustment for eGFR may partly adjust away the exposure of interest, and the null estimate in Model 4 should be interpreted with this in mind. Fifth, the exploratory MIMIC-IV analysis was based on administrative diagnosis codes, included few stone cases, and was not designed as external validation, so it can only indicate the direction of an association. Finally, the observational design leaves the study susceptible to residual confounding by unmeasured factors, and the machine-learning results are descriptive and should not be used for clinical prediction.

## 5. Conclusions

In this large, nationally representative sample of US adults, higher BAR was associated with kidney stones in models that did not account for renal function and metabolic status, but the association did not persist after adjustment for BMI, diabetes, eGFR and PIR, and no nonlinear threshold was retained after adjustment. BAR should therefore be regarded as a marker that aggregates renal and metabolic risk rather than as an independent risk factor for kidney stones. It does not appear to add information beyond established metabolic and renal measures, and it should not be used as a stand- alone risk marker. Personalized precision prevention strategies for kidney stones should focus on the modifiable metabolic factors identified here, including obesity and diabetes, rather than on BAR itself.

### Declarations

#### Ethics approval and consent to participate

The National Health and Nutrition Examination Survey (NHANES) is a publicly available database conducted by the National Center for Health Statistics (NCHS) of the U.S. Centers for Disease Control and Prevention (CDC). The NHANES study protocol was reviewed and approved by the NCHS Research Ethics Review Board (ERB) (Protocol #2005-06, #2011-17, and #2018-01). All participants provided written informed consent prior to their enrollment in the original survey. The present study is a secondary analysis of de-identified, publicly available NHANES data. According to U.S. federal regulations under 45 CFR 46.104(d)(7) (the Common Rule), research involving publicly available, de-identified data is exempt from additional institutional review board approval. This secondary analysis was conducted in accordance with the ethical principles of the Declaration of Helsinki.

## Consent for publication

Not applicable. No identifying images or other personal or clinical details of participants are presented in this manuscript.

## Availability of data and materials

The datasets analyzed in this study are publicly available from the NHANES website: https://www.cdc.gov/nchs/nhanes. MIMIC-IV data are available from PhysioNet to credentialed users (https://physionet.org/content/mimiciv/).

## Author contributions

**Fu Zhu:** Methodology, Software, Formal analysis, Visualization, Writing-original draft.

**Chao Wang and Yujie Wang: Data curation, Validation.**

**Jianqiang Bian and Dongpeng Zhang: Conceptualization, Supervision, Writing – review & editing.**

All authors read and approved the final manuscript.

Competing interests: The authors declare that they have no competing interests.

## Funding

Not applicable

## Data Availability

The data used in this study are publicly available from the National Health and Nutrition Examination Survey (NHANES), conducted by the National Center for Health Statistics (NCHS), Centers for Disease Control and Prevention (CDC). All NHANES datasets are freely accessible without restriction at https://www.cdc.gov/nchs/nhanes/. No new data were generated in this study.

## Acknowledgements

Not applicable

## Notes

### Competing Interest Statement

The authors have declared no competing interest.

### Author Declarations

The National Health and Nutrition Examination Survey (NHANES) protocols were approved by the NCHS Research Ethics Review Board (ERB) of the National Center for Health Statistics, Centers for Disease Control and Prevention. This study is a secondary analysis of publicly available, de-identified NHANES data and was therefore exempt from additional institutional review board approval.

### Summary of Updates

The analytic dataset was rebuilt from the NHANES 2007 to 2020 cycles using the current data release. The sample size changed from 22,874 to 23,652 participants, and four covariates were added to the adjustment set: body mass index, diabetes, estimated glomerular filtration rate and poverty income ratio. The revised analysis presents a four model adjustment ladder and designates the fully adjusted model as the primary analysis. These changes altered the main results. The association between the blood urea nitrogen to albumin ratio and kidney stones was attenuated to the null after full adjustment. The Abstract, Results, Discussion and Conclusions were rewritten around this null finding, and the statements that the ratio could serve as an independent risk factor or as a risk stratification tool were removed. The revised manuscript reports a sensitivity analysis restricted to participants with preserved kidney function, an incremental adjustment analysis, restricted cubic spline and XGBoost analyses, and an exploratory analysis in the MIMIC IV cohort. Tables 1 to 6, supplementary Tables S1 to S4 and Figures 1 to 7 were updated, corrected or added accordingly, and several figure panels were redrawn. The reference list was checked against the cited text, reduced from 43 to 39 entries and reformatted in Vancouver style. The title page, author details, ethics statement and declarations were corrected.

